# Inclusion of cycle threshold (CT) values when reporting SARS-CoV-2 RT-PCR results improves clinical Interpretation in suspected and confirmed COVID-19

**DOI:** 10.1101/2021.02.11.21251557

**Authors:** Peter V. Coyle, Naema Hassan Al Molawi, Mohamed Ali Ben Hadj Kacem, Reham Awni El Kahlout, Einas Al Kuwari, Abdullatif Al Khal, Imtiaz Gilliani, Andrew Jeremijenko, Hatoun Saeb, Sheikh Mohammad Al Thani, Roberto Bertollini, Hanan F. Abdul Rahim, Hiam Chemaitelly, Patrick Tang, Ali Nizar Latif, Saad Al Kaabi, Muna A. Rahman S. AlMaslamani, Brendan David Morris, Nasser Al-Ansari, Anvar Hassan Kaleeckal, Laith J. Abu Raddad

**Author notes:** Corresponding author Study approved by Hamad Medical Corporation Institutional Review Board.

## Abstract

**Introduction:** The Cycle Threshold (CT) value in Real-time Polymerase Chain Reaction (RT-PCR) is where a target specific amplification signal becomes detectable and can infer viral load, risk of transmission and recovery in SARS-CoV-2 infections. Adoption into routine practice is however uncommon.

**Gap Statement:** The lack of inclusion of CT values when reporting SARS-CoV-2 RT-PCR results in routine practice.

**Aim:** To use CT values when reporting SARS-CoV-2 RT-PCR results in Qatar to aid clinical interpretation and patient management.

**Methodology:** Routine CT values across 3 different RT-PCR platforms were reviewed for concordance at presentation and clearance in patients with COVID-19. An Indicative Threshold of CT 30 based on viral clearance kinetics categorized low and high CT values.

**Results:** There was very high Correlation and Kappa Score agreement between the different gene targets in each platform (p<0.001). Using the Indicative Threshold it was possible to autoverify and add average CT values and append Interpretive Comments to all RT-PCR reports. The new reporting algorithm impacted immediately and safely on: physician interpretation of SARS-CoV-2 results; patient management; staff surveillance protocols; length of stay in quarantine; a redefinition of patient recovery.

**Conclusion:** Incorporation of CT values into routine practice is possible across different RT-PCR platforms and adds useful information for patient management. The use of an Indicative Threshold and interpretive comments improves clinical interpretation of the result and could be a model for reporting other respiratory infections. The current accepted practice of withholding CT values should be reviewed by the profession, accreditation bodies and regulators.

## Introduction

The World Health Organization (WHO) reported: clusters of viral pneumonia on December 31, 2019 in Wuhan China; isolation of SARS-CoV-2 on January 9, 2020; definition of Coronavirus Infectious Disease 2019 (COVID-19) caused by the virus; a Public Health Emergency of International Concern on January 30, 2020; a pandemic on March 11, 2020 [1]. WHO advised the use of Reverse Transcriptase Polymerase Chain Reaction (RT-PCR) on combined nasopharyngeal / oropharyngeal swabs (NPS/OPS) for diagnosis of COVID-19 and two negative RT-PCRs 24 hours apart for defining recovery[2].

RT-PCR amplifies over 40 to 45 cycles, with the cycle threshold (CT) value being determined where the fluorescent signal of a test exceeds the background signal of controls. The CT value is inverse to the viral load, with clearance indicated by rising CTs. This is because more PCR cycles are required to detect the virus. Using CTs in estimating viral load and transmission risk is not routine because of perceived lack of platform to platform concordance and the absence of a direct link between CT values and clinical outcome.

A positive result for SARS-CoV-2 is defined by the manufacturer-specified CT values for one or more genes, with an inconclusive result reported where the combination of reactive genes fails to meet the definition.

After infection SARS-CoV-2 has a short latent period where virus is undetectable for about 4 days, followed by a period of high viral load coinciding with the pre-symptomatic and symptomatic transmission phase, lasting for another four days, and then followed by prolonged viral RNA shedding [3-8]. The timing of transmission varies with the data source with Ganyani reporting the generation interval (time between infection events in an infector-infectee pair) as 5.20 days (95%: CrL 3.78–6.78) and 3.95 days (95% CrI: 3.01–4.91) for two separate outbreaks [9].

At the onset of symptoms patients present with high viral loads (<CT 20) which subsequently fall in both upper and lower respiratory tracts, with higher loads in lower airways than those seen in NPS/OPS specimens [10]. Detection of low level viral RNA can last for weeks to months but transmission does not typically exceed 10 days from first PCR or symptom onset in mild infections and 20 days in severe illness [10-12]. Prolonged shedding and categorical reporting (positive or negative) can risk inferring patients are infectious when they are in recovery, leading often to unnecessary extension of quarantine.

Attempts to use CT values as a surrogate of viral load and by extrapolation of disease severity have been reported but not widely adopted. Wishaupt [13] showed low CT values in common respiratory viral infections, including seasonal human coronaviruses (HCoV), were associated with hospitalisation, increased oxygen requirement and longer lengths of hospital stay. Spencer categorised influenza infections on the basis of an arbitrarily set CT threshold, separating them into low and high CT values to differentiate high and low viral loads respectively [14]. They confirmed the low CT cohort was seen primarily in younger patients (3 – 8y), symptomatic elderly patients and presentation within two days of symptom onset, indicating early acute infection. A similar pattern of CT values was reported in children with *Bordetella pertussis* with low values indicating high bacterial loads in unvaccinated children with life threatening infections; bacterial loads were three orders of magnitude higher than those seen in vaccinated children [15].

CT values have also been used to estimate the period of infectivity in COVID-19. Bullard [11] investigated virus recovery from symptom onset and found that virus could not be isolated after 8 days or CT values > 24. Centers for Disease Control and Prevention (CDC) [16] and La Scola [12] reported virus recovery was restricted to the first 8 days of a symptomatic infection and CT values <33. Wolfe [10] and van Kampen [17] found viral recovery correlated with high viral loads and by implication low CT values, with the observation that viral recovery in patients with severe disease could be prolonged [17]. Time based prediction of infectiousness based on virus culture is more consistent than individual CT thresholds although likely to overestimate the period of infectivity for most people.

Adoption of CT value reporting into routine practice faces hurdles associated with regulatory bodies and accreditation requirements that look for precise validation of clinical interpretation of individual CT values on each platforms. However not reporting CT values deprives physicians and patients of valuable information for interpreting the meaning of a positive result. In the current study three result categories were reported based on manufacturer guidance from the respective CT values of each gene target: positive, negative and inconclusive. A follow-up sample was requested where an inconclusive test result was reported, and CT values were not included.

To meet test demand in Qatar with a population of 2.8 million, three platforms -Roche, Thermo-Fisher and Cepheid, respectively amplifying 2, 3 and 2 gene targets, were used for SARS-CoV-2 RT-PCR. A review of their respective assay performance was undertaken to see whether CT values could be safely added to reporting and improve interpretation of SARS-CoV-2 results.

## Methods

### RT-PCR Assay Platforms

All assays were pre-validated before use in line with the College of American Pathologists accreditation standards. Each platform and reagent combination was analysed for comparative amplification kinetics of the respective gene targets. The systems in use prior to the introduction of CT reporting were: (a) Automated Platforms -the Roche cobas® 6800 system using the cobas® SARS-CoV-2 Test targeting the E and orf1a/b genes (Roche, Switzerland) and the Xpert® Xpress SARS-CoV-2 targeting the E and N genes (Cepheid, USA); (b) Manual platforms -EZ1 (QIAGEN, USA) and QIAsymphony (QIAGEN, USA) extraction with thermal cycling using TaqPath™ PCR COVID-19 Combo Kit targeting the N, S and orf1a/b genes (Thermo Fisher Scientific, USA) on ABI 7500 thermal cyclers (Thermo Fisher Scientific, USA).

### Thermal Cycling CT Kinetics

A total of 173,557 individual CT values on 148,066 infected persons across the Roche, Thermo-Fisher and Cepheid platforms were analysed for concordance using routine CT values from the respective instruments. Sequential samples on the Roche and Thermo Fisher platforms were also reviewed to establish the viral kinetics associated with viral clearance during recovery.

### Indicative Threshold For Reporting CT Values

By reference to observed clearance on the respective platforms and publications linked to reduced transmission, a threshold below and above which transmission was more or less likely was established. The time taken to reach average CT values from initial infection to viral clearance was reviewed and the average CT values at day 14 was taken as a potential Indicative Threshold (IT) associated with reduced transmission.

### Reporting Change

Reporting was redesigned with reference to the Indicative Threshold. Individual gene CT values with their alphanumeric results were run through a flat file interface to the mainframe computer. An average CT value was automatically calculated, and the resulting categories added based on both the average and individual CT values. Four result categories were used: 1) positive 2) reactive, (CT>30); 3) negative; 4) inconclusive. An interpretive comments table was designed and automatically appended at auto-verification. The specimen type with individual tracking times at each stage from collection to reporting was used to calculate turnaround times each day. The four reported categories were also interfaced in real time to the National Dashboard, for daily review, and were used to trigger a colour change on Qatar’s national COVID-19 track and trace EHTERAZ application.

### Follow-up Audit of Enhanced Reporting

Viral CTs in positive samples within four days from a reactive, inconclusive or negative result were reviewed and plotted as arbitrary log values (CT40 – CT value)/3.3) against the day of becoming positive. Cross referencing the four result categories against a seroprevalence research database was also used to determine seroprevalence in each category -Table 1. Two categories of positive result were cross referenced: those with one or > one RT-PCR positive result.

**Table 1.** Specimen numbers per RT-PCR reporting category and seroprevalence database

| Reporting Category | Specimen Number | Period Covered |
| --- | --- | --- |
| Positive | 116213 | 02/04/2020 - 25/08/2020 |
| Reactive | 11291 | 24/06/2020 - 25/08/2020 |
| Inconclusive | 32678 | 08/03/2020 - 02/09/2020 |
| Negative | 745673 | 04/03/2020 - 29/08/2020 |
| Seroprevalence | 149878 | 28/06/2020 -23/08/2020 |

### Statistics

Spearman Rank Coefficient and Kappa scores were used for comparison of CT values across each platform. Box-plots of viral kinetics with median and Inter-Quartile ranges were used in follow-up positive samples from Day 1 to Day 4 following a negative result.

## Results

### CT Kinetics across Thermal Cycling

The assay performance during the period of the study was acceptable in internal and external proficiency panel assessments in line with the College of American Pathologists accredited Quality Assurance program of the laboratory.

The CT values across the different gene targets for each of the three platforms showed good agreement as measured by Spearman correlation coefficient and Kappa scores – Table 2, in keeping with acceptable performance during routine practice.

**Table 2.**
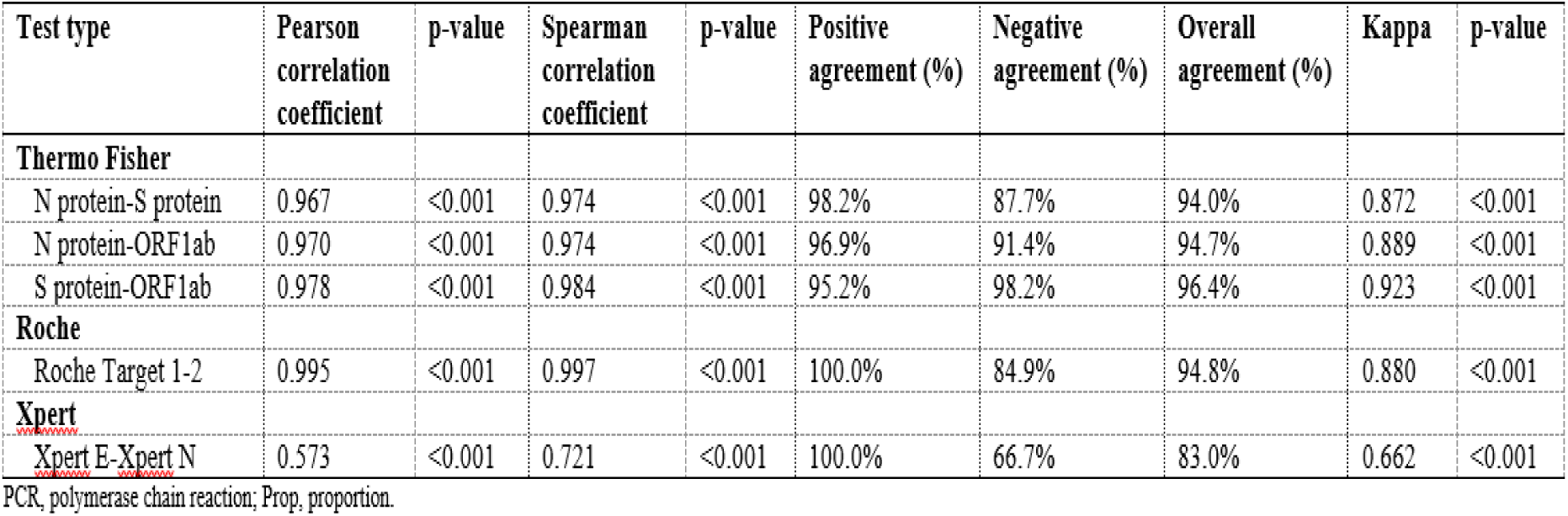
Correlation of Gene Targets Across The Roche, Thermo Fisher and Cepheid Platforms

| Test type | Pearson correlation coefficient | p-value | Spearman correlation coefficient | p-value | Positive agreement (%) | Negative agreement (%) | Overall agreement (%) | Kappa | p-value |
| --- | --- | --- | --- | --- | --- | --- | --- | --- | --- |
| **Thermo Fisher** |  |  |  |  |  |  |  |  |  |
| N protein-S protein | 0.967 | <0.001 | 0.974 | <0.001 | 98.2% | 87.7% | 94.0% | 0.872 | <0.001 |
| N protein-ORF1ab | 0.970 | <0.001 | 0.974 | <0.001 | 96.9% | 91.4% | 94.7% | 0.889 | <0.001 |
| S protein-ORF1ab | 0.978 | <0.001 | 0.984 | <0.001 | 95.2% | 98.2% | 96.4% | 0.923 | <0.001 |
| **Roche** |  |  |  |  |  |  |  |  |  |
| Roche Target 1-2 | 0.995 | <0.001 | 0.997 | <0.001 | 100.0% | 84.9% | 94.8% | 0.880 | <0.001 |
| **Xpert** |  |  |  |  |  |  |  |  |  |
| Xpert E-Xpert N | 0.573 | <0.001 | 0.721 | <0.001 | 100.0% | 66.7% | 83.0% | 0.662 | <0.001 |

PCR, polymerase chain reaction; Prop, proportion.

The Roche and Thermo Fisher assays confirmed an early decline in viral load, as indicated by rising CT values across each platform, followed by a prolonged period of low level detection of viral RNA -Figure 1. The rise in CT values observed during the recovery phase was similar for each gene target, with the Roche assay detecting more prolonged shedding at a higher CT level than the Thermo Fisher assay.

**Figure 1.**
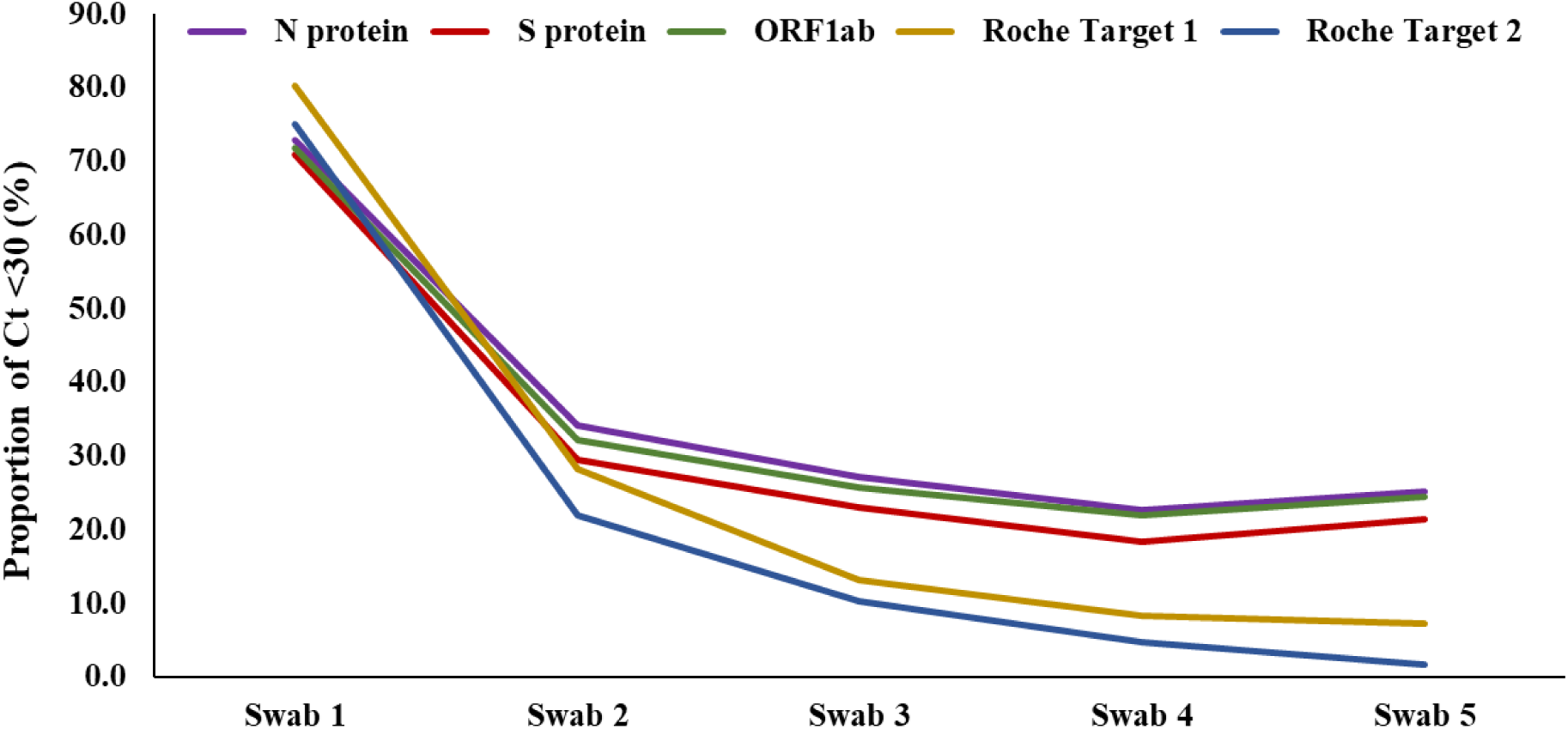
Progression of proportion of CT <30 over number of swabs tested

### Indicative CT Threshold

Taking the lowest CT value as indicating the start of detectable virus by RT-PCR, and assuming that infected persons spend on average a total PCR positivity duration of four weeks (28 days), as informed by current evidence [18, 19], Figure 2 shows the number of days spent at or below each CT value across the different platforms; a CT 28 value equated to Day 14. The Indicative Threshold selected for defining the new reactive reporting category was set at CT>30 to arbitrarily build in an additional margin where the likelihood of transmission was deemed low and being in recovery high for most recovering patients.

**Figure 2.**
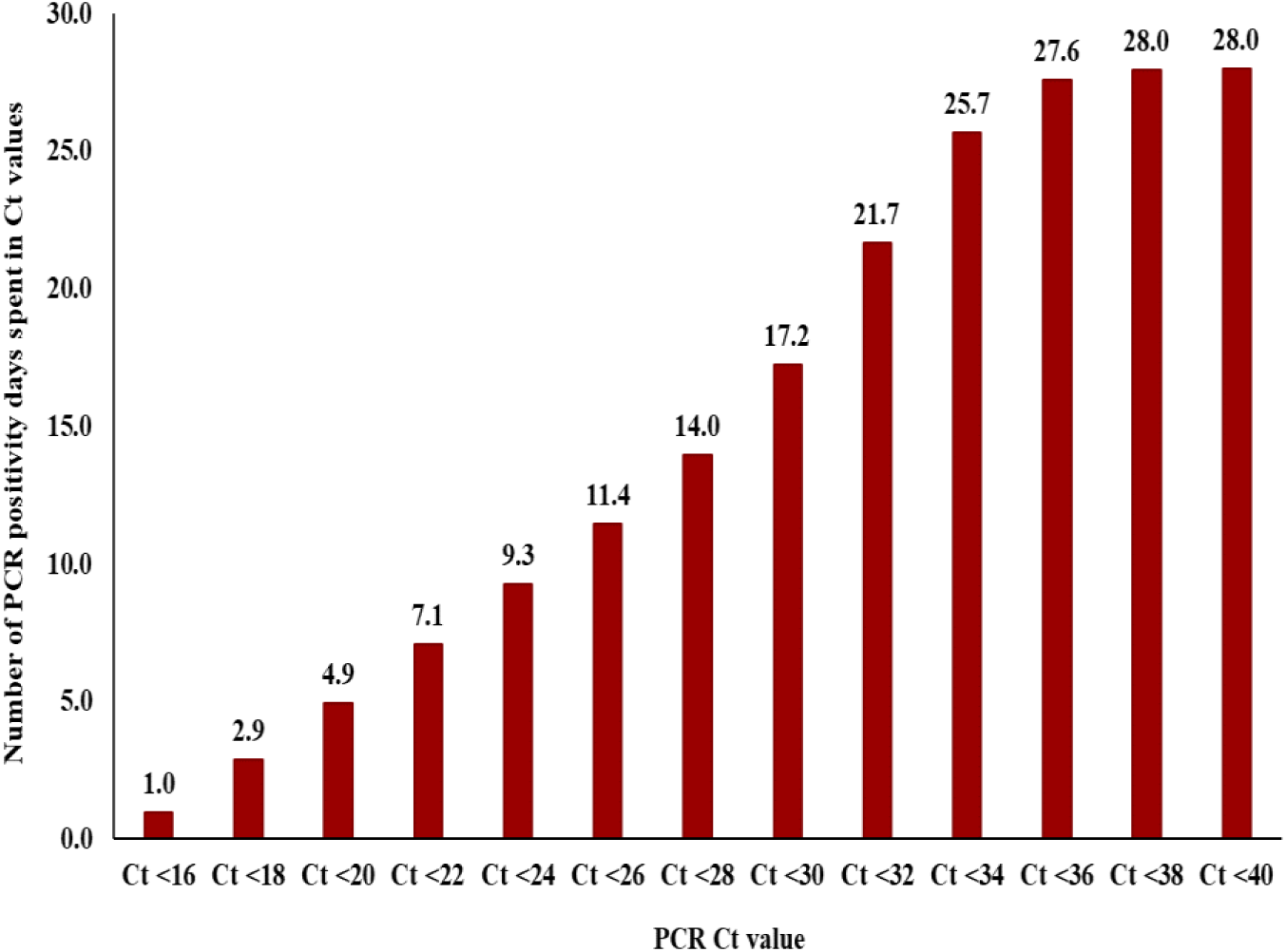
RT-PCR positivity days by Cycle Threshold value

### Reporting using the Indicative Threshold

The use of an Indicative Threshold of CT>30 allowed the use of 4 reporting categories. Interpretive Comments based on the kinetics of SARS-CoV-2 replication and likely COVID-19 clinical presentation were added at autoverification of results, Table 3. Comments emphasised the need for repeat testing in an outbreak and the need for clinical assessment of patients irrespective of the CT values.

**Table 3.** Interpretive Comments for Enhanced Reporting

| **Interpretive Comments for NPS/OPS Swab Results** |  |  |  |
| --- | --- | --- | --- |
| **Positive (CT ≤30)** | **Reactive (CT >30)** | **Negative** | **Inconclusive** |
| SARS-CoV-2 RNA detected – regarded as potentially infectious up to 10 days from symptom onset or 1st PCR positive; prolonged shedding seen in severe cases | In asymptomatic patients usually seen in recovery phase which can be prolonged. | SARS-CoV-2 virus RNA NOT detected – excludes COVID-19. | In asymptomatic patients usually seen in recovery phase which can be prolonged. Repeat if detected in pre-procedure screening. |
| Low and high CT values indicate high and low viral loads respectively | Repeat where clinical suspicion remains / in an outbreak / after contact with a case. | Repeat where clinical suspicion remains / in an outbreak / after contact with a case | Repeat where clinical suspicion remains / in an outbreak / after contact with a case |
| Clinical assessment essential regardless of CT value | Clinical assessment essential regardless of CT value | Clinical assessment essential regardless of CT value | Clinical assessment essential regardless of CT value |

### Confirmation of COVID-19 after Follow-up of RT-PCR Negative Samples

In review of patients initially reported with non-positive result (reactive, inconclusive or negative) it was found that a positive (CT≤30) result was reported within 10 days in 5.2%, 1.9% and 7.1% of reactive, negative and inconclusive results respectively; acute early infections, defined by a CT<20, were reported in 0.2%, 0.8% and 1.6% of cases – Table 4.

**Table 4.** Follow-up of patients initially reported reactive, inconclusive or negative

| Category | CT<20 | CT<30 |
| --- | --- | --- |
| Reactive | 26 / 11291 (0.2%) | 591 / 11291 (5.2%) |
| Negative | 2515 / 328095 (0.8%) | 6429 / 328095 (1.9%) |
| Inconclusive | 460 / 29104 (1.6%) | 2066 / 29104 (7.1%) |

Within 4 days of a negative result a rapid increase in load was seen on day 1 which continued to rise on days 2 and 3 in keeping with the predicted transition from the asymptomatic to a symptomatic phase of infection – Figure 3 [3].

**Figure 3.**
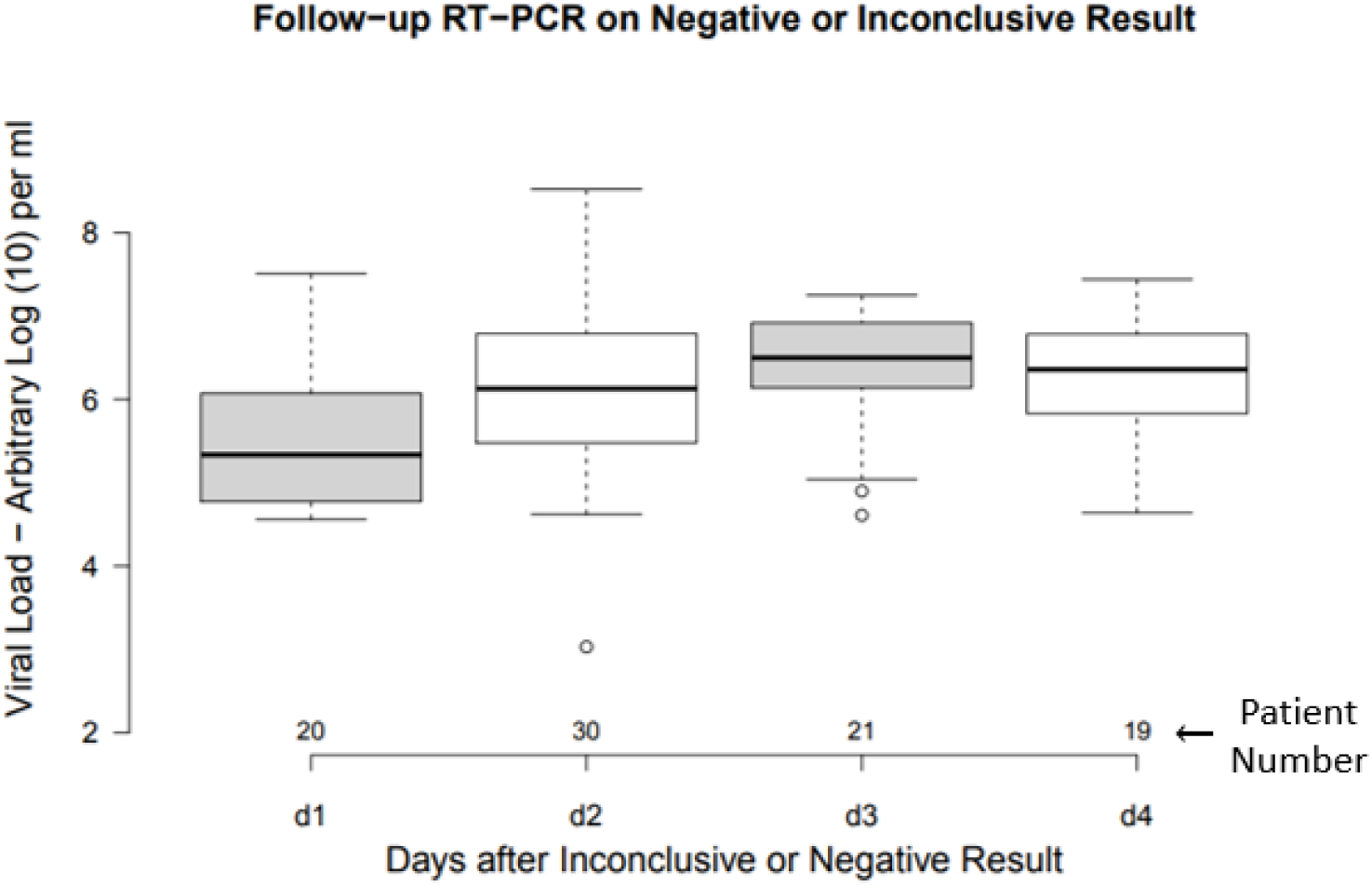
Transition from Non-detectable to Detectable After Negative or Inconclusive Results

### Seroprevalence by Reporting Category

The respective seroprevalence of patients in the different Reporting Categories when cross-referenced against the Seroprevalence database is shown in Figure 4. There was a trend to higher seroprevalence respectively for negative, single positive RT-PCR, inconclusive, >1 positive RT-PCR and reactive.

**Figure 4.**
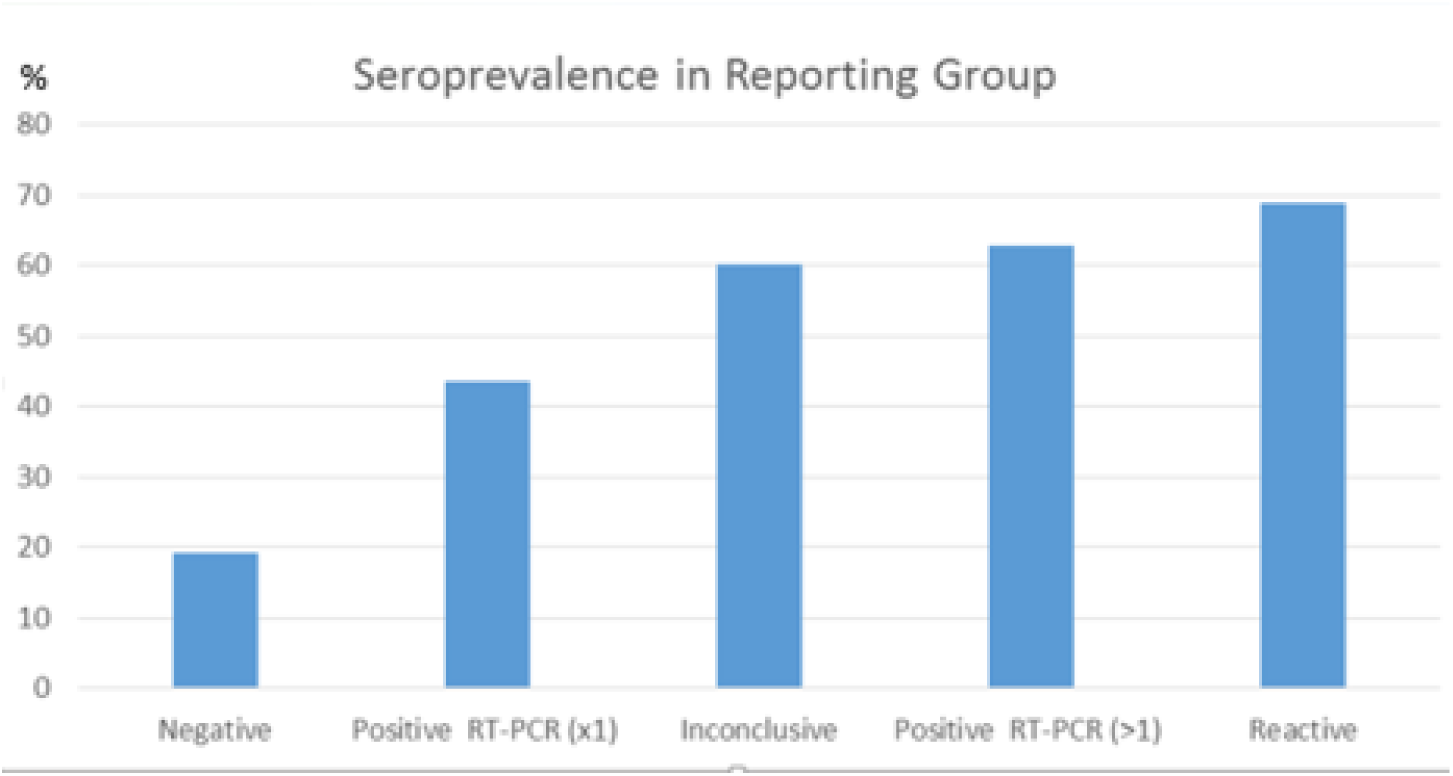
Seroprevalence associated with Reporting Category

## Discussion

The decision to include CT values in routine reporting of COVID-19 in Qatar was felt to be possible following a review of the respective test platforms and was communicated in advance to physicians and Public Health teams dealing with the outbreak. While exclusion of CT values is normal practice when reporting RT-PCT results, the implications of a SARS-CoV-2 result are critical to clinical management and Public Health decisions. A confirmed infection can present over a 10 Log_10_ range, making categorical reporting of results as positive or negative restrictive and open to misinterpretation. Reporting of CT values with interpretive comments helped increased the interpretation and safety of reporting and could be modified to reflect changes in the clinical knowledge base of the condition.

Not reporting of CT values stems from assumed lack of platform to platform concordance and lack of specific outcomes linked to individual CT values [20]. The approach described here does not define absolute thresholds or pre-set CT values for clinical interpretation. The Indicative Threshold categorised results around a CT value based on the viral kinetics of clearance. This along with the interpretive comments allowed inference of the phase of infection and potential for continuing risk of transmission. Since most RT-PCR platforms provide similar amplification profiles, as demonstrated in Figure 2, the use of an Indicative Threshold is a flexible way to allow reporting across platforms. Correlation of new with existing platforms can be formally undertaken and this is now part of our routine pre-use validation.

The approach described could be applied to other respiratory infections. Curran reported CT values for *Bordetella pertussis* where low CT values were seen in unvaccinated children with Whooping Cough compared to those who had received the vaccine (mean CT 23 versus mean CT 33; p > 0.001); Interpretive Comments were not reported [15]. These low CT *Bordetella* infection in pre-vaccinated children gives rise to potentially life threatening infections which are associated with a significant transmission risk to unvaccinated staff and family.

While the seroprevalence results in Figure 4 indicated that most Reactive and Inconclusive reports were in keeping with a previous SARS-CoV-2 infection rather than a false positive result, in a small number of cases they indicated acute early infection – Table 4. Here ninety patients with confirmed infection immediately following a negative result showed a rapid increase in viral load in-keeping with a transition from latent to promiscuous replication as described in the predictive model of Li [3]. Reporting CT values in these situations provides clinicians with additional information for informing their differential diagnosis.

The introduction of CT reporting saw a fall in enquiries to the specialist COVID-19 advisory call centre that had been established in Qatar for COVID-19 (John Mitchel personal communication). It also led to a reduction in enquiries from front line clinical staff to virology (in the author’s personal experience). In addition a group of microbiology consultants providing interpretation advice to clinical teams, was considered no longer required, and they were able to return to routine duties.

For patient admission and discharge policy the new reporting provided an objective categorisation for assisting patient flow that was used for admission, transfer and discharge from COVID and non-COVID designated hospitals respectively. This increased flexibility improved the efficiency of a sometimes over stretched service.

Use of CT values also helped triage arrangements for quarantine. WHO guidelines for 2 negative RT-PCR results 24 hours apart often extended the period of quarantine for confirmed infection beyond 14 days. This occurred because of the prolonged shedding seen in COVID-19 and a reluctance to discharge patients with a positive result[2]. Reporting CT values with Interpretive comments linked to the CDC time-based discontinuation of transmission precautions allowed a more coherent discharge policy. While CDC guidelines advocated a 10 day period from symptoms or first positive PCR [21], the Indicative Threshold was aligned to a more precautionary 14 day estimate. These arrangements provided a better definition of recovery and as a result the numbers of recovered patients rose steeply.

The need for and duration of quarantine for patients with a Reactive result was reduced from 14 to 7 days as these patients were deemed suitable for home self-isolation. This new policy reduced the numbers in hotel or hospital quarantine and allowed individuals and families a significant financial saving and better well-being by avoiding or reducing the need for hotel quarantine accommodation that was introduced in response to the pandemic.

The policy also supported the country’s tracking and tracing application EHTERAZ (meaning PRECAUTION) which is a contact tracing application installed on mobile devices used by individuals when leaving their homes. EHTERAZ defines 4 categories of health status by colour: Green – no current positive result; Yellow – in quarantine; Grey – suspected or exposed; Red – currently positive. The new reactive category allowed release of people from quarantine to home isolation with a change of status from Red to Yellow. From a safety perspective this was also supported by a report from the Korean Centers for Disease Control that confirmed patients with on-going positive RT-PCR were not a transmission risk [22].

### Conclusion

The use of CT values and interpretive comments in routine reporting improved understanding of SARS-CoV-2 results and impacted on patient management and Public Health delivery. The current accepted practice of withholding CT values should be reviewed by the profession, accreditation bodies and regulators.

## Data Availability

All relevant data are available within the manuscript.

## References

1. World Health Organization: Timeline of WHO’s response to COVID-19. In., vol. https://www.who.int/news/item/29-06-2020-covidtimeline; 2020: Accessed on 9 September 2020; https://www.who.int/news/item/2029-2006-2020-covidtimeline.

2. World Health Organization: Laboratory testing of 2019 novel coronavirus (2019-nCoV)in suspected human cases: interim guidance, 17 January 2020. Geneva: World Health Organization; 2020.

3. Li R, Pei S, Chen B, Song Y, Zhang T, Yang W, Shaman J: Substantial undocumented infection facilitates the rapid dissemination of novel coronavirus (SARS-CoV2). Science 2020, 368(6490):489–493.

4. Lauer SA, Grantz KH, Bi Q, Jones FK, Zheng Q, Meredith HR, Azman AS, Reich NG, Lessler J: The Incubation Period of Coronavirus Disease 2019 (COVID-19) From Publicly Reported Confirmed Cases: Estimation and Application. Annals of internal medicine 2020, 172(9):577–582.

5. World Health Organization: Report of the WHO-China Joint Mission on Coronavirus Disease 2019 (COVID-19). Available from :https://www.who.int/docs/default-source/coronaviruse/who-china-joint-mission-on-covid-19-final-report.pdf. Accessed on March 10, 2020. In.; 2020.

6. Rothe C, Schunk M, Sothmann P, Bretzel G, Froeschl G, Wallrauch C, Zimmer T, Thiel V, Janke C, Guggemos W et al: Transmission of 2019-nCoV Infection from an Asymptomatic Contact in Germany. The New England journal of medicine 2020, 382(10):970–971.

7. Zou L, Ruan F, Huang M, Liang L, Huang H, Hong Z, Yu J, Kang M, Song Y, Xia J et al: SARS-CoV-2 Viral Load in Upper Respiratory Specimens of Infected Patients. The New England journal of medicine 2020, 382(12):1177–1179.

8. He X, Lau EHY, Wu P, Deng X, Wang J, Hao X, Lau YC, Wong JY, Guan Y, Tan X et al: Temporal dynamics in viral shedding and transmissibility of COVID-19. Nature medicine 2020, 26(5):672–675.

9. Ganyani T, Kremer C, Chen D, Torneri A, Faes C, Wallinga J, Hens N: Estimating the generation interval for coronavirus disease (COVID-19) based on symptom onset data, March 2020. Euro surveillance : bulletin Europeen sur les maladies transmissibles = European communicable disease bulletin 2020, 25(17):doi:10.2807/1560-7917.Es.2020.2825.2817.2000257.

10. Wölfel R, Corman VM, Guggemos W, Seilmaier M, Zange S, Müller MA, Niemeyer D, Jones TC, Vollmar P, Rothe C et al: Virological assessment of hospitalized patients with COVID-2019. Nature 2020, 581(7809):465–469.

11. Bullard J, Dust K, Funk D, Strong JE, Alexander D, Garnett L, Boodman C, Bello A, Hedley A, Schiffman Z et al: Predicting infectious SARS-CoV-2 from diagnostic samples. Clinical infectious diseases : an official publication of the Infectious Diseases Society of America 2020, doi:10.1093/cid/ciaa638: Epub ahead of print.

12. La Scola B, Le Bideau M, Andreani J, Hoang VT, Grimaldier C, Colson P, Gautret P, Raoult D: Viral RNA load as determined by cell culture as a management tool for discharge of SARS-CoV-2 patients from infectious disease wards. European journal of clinical microbiology & infectious diseases : official publication of the European Society of Clinical Microbiology 2020, 39(6):1059–1061.

13. Wishaupt JO, Ploeg TV, Smeets LC, Groot R, Versteegh FG, Hartwig NG: Pitfalls in interpretation of CT-values of RT-PCR in children with acute respiratory tract infections. Journal of clinical virology : the official publication of the Pan American Society for Clinical Virology 2017, 90:1–6.

14. Spencer S, Chung J, Thompson M, Piedra PA, Jewell A, Avadhanula V, Mei M, Jackson ML, Meece J, Sundaram M et al: Factors associated with real-time RT-PCR cycle threshold values among medically attended influenza episodes. Journal of medical virology 2016, 88(4):719–723.

15. Curran T, Coyle PV: Understanding the true burden and infection dynamics of Bordetella pertussis using molecular diagnostics. The Journal of infection 2016, 72(4):504–505.

16. Centers for Disease Control and Prevention: Duration of Isolation and Precautions for Adults with COVID-19. In.; 2020: Accessed September 2020: https://www.cdc.gov/coronavirus/2019-ncov/hcp/duration-isolation.html.

17. van Kampen JJA, van de Vijver DAMC, Fraaij PLA, Haagmans BL, Lamers MM, Okba N, van den Akker JPC, Endeman H, Gommers DAMPJ, Cornelissen JJ et al: Shedding of infectious virus in hospitalized patients with coronavirus disease-2019 (COVID-19): duration and key determinants. medRxiv 2020:2020.2006.2008.20125310.

18. Sethuraman N, Jeremiah SS, Ryo A: Interpreting Diagnostic Tests for SARS-CoV-2. JAMA 2020.

19. Wajnberg A, Mansour M, Leven E, Bouvier NM, Patel G, Firpo A, Mendu R, Jhang J, Arinsburg S, Gitman M et al: Humoral immune response and prolonged PCR positivity in a cohort of 1343 SARS-CoV 2 patients in the New York City region. medRxiv 2020:2020.2004.2030.20085613.

20. Public Health England: Cycle threshold (Ct) in SARS-CoV-2 RT-PCR: A guide for health protection teams about interpreting Ct in real-time, reverse transcription polymerase chain reaction (RT-PCR) assays performed for Sars-CoV-2. In.; 2020: Accessed 28 October 2020 : https://www.gov.uk/government/publications/cycle-threshold-ct-in-sars-cov-2022-rt-pcr.

21. Centers for Disease Control and Prevention: Duration of Isolation and Precautions for Adults with COVID-19. In.; 2020: Accessed 19 October 2020: https://www.cdc.gov/coronavirus/2019-ncov/hcp/duration-isolation.html.

22. Korea Disease Control and Prevention Agency: Findings from investigation and analysis of re-positive cases. In.: Korea Disease Control and Prevention Agency; 2020: Accessed 19 May 2020: http://www.kdca.go.kr/board/board.es?mid=a30402000000&bid=30402000030.

